# Assessing the influence of parental anxiety on childhood anxiety during the COVID-19 pandemic in the United Arab Emirates

**DOI:** 10.1101/2020.06.11.20128371

**Authors:** Basema Saddik, Amal Hussein, Ammar Albanna, Iffat Elbarazi, Arwa Al-Shujairi, Fatemeh Saheb Sharif-Askari, Mohamad-Hani Temsah, Emmanuel Stip, Qutayba Hamid, Rabih Halwani

## Abstract

The COVID-19 pandemic originated in Wuhan, China on December 31^st^ and spread into international borders, leading to a public health crisis and complete shutdown of countries. The strict quarantine measures taken by governments kept a large number of people, around the world, in isolation and affected many aspects of people’s lives. These unprecedented changes triggered a wide variety of psychological problems ranging from panic disorders, anxiety and depression. In this study, we aim to explore anxiety levels among parents, teachers and the general community amid the COVID-19 pandemic in the UAE, as well as identify emotional and anxiety disorders in children. Using a web-based cross-sectional survey we collected data from 2,200 self-selected assessed volunteers. Demographic information, knowledge and beliefs about COVID-19, generalized anxiety disorder (GAD) using the (GAD-7) scale, emotional problems in children using the strengths and difficulties questionnaire (SDQ), worry and fear about COVID-19, coping mechanisms and general health information were collected. The overall prevalence of GAD in the general population was 71% with younger people (59.8%) and females (51.7%) reporting the highest levels of anxiety. Parents who were teachers reported the highest percentage of emotional problems in children (26.7%) compared to parents only (14.6%) or teachers only (4.7%). Multivariate logistic regression for GAD-7 score showed that females, participants who felt public fear was justifiable, persons who worried about COVID-19, persons who intended to take the COVID-19 vaccine and smokers were all associated with anxiety. Multivariate logistic regression for SDQ showed parents who had severe anxiety levels were 7 times more likely to report more emotional problems in their children (OR=7.00, 95% CI, 3.45 to 14.0) than less anxious parents. Findings suggest the urgency of policy makers to develop effective screening and coping strategies for parents and teachers and more specifically for vulnerable children.

## 1. Introduction

The Coronavirus Disease 2019 (COVID-19) outbreak emerged in Wuhan, China, in December 2019 and was declared a public health emergency on January 30^th^ [1] and subsequently a global pandemic by the World Health Organization (WHO) on March 11^th^ 2020 [2]. As of May 31^st^ 2020, more than 6 million laboratory-confirmed cases and over 370,000 deaths were reported worldwide, with 33,896 confirmed cases and 262 deaths in the United Arab Emirates (UAE) [3]. In the absence of an effective treatment or vaccine for this infectious disease, unprecedented public health interventions were implemented across the UAE and countries around the world to curb the transmission of this life-threatening disease. These included the closure of international borders, travel bans, government-enforced lockdowns and isolations, closure of schools and academic institutions, strict social, and physical distancing and quarantine. These precautionary measures, along with the uncertainty and fear of an emerging pandemic and the unusual disruption to the way people live, work and study, are likely to have significant and long-term implications on the mental health of a community [4].

Research on past infectious disease outbreaks, such as severe acute respiratory syndrome (SARS), swine flu, influenza,and Ebola, revealed a wide range of psychosocial impacts at the individual, community, and international levels. These included worries about contracting infections and fear of dying [5]; the significant increase of psychiatric conditions including anxiety disorders, post-traumatic stress disorder and depressive disorders [6]; feelings of helplessness and panic and increased perception of risk [7-9]. More recently, studies investigating the psychological impacts of the COVID-19 outbreak in China have reported moderate to severe stress, generalized anxiety disorder, insomnia, and depression in the general population [10, 11].

However, despite the number of studies that have investigated the psychosocial impact of infectious disease outbreaks in adults, there remains limited information about the mental health impact of the COVID-19 pandemic in the general population, and there are no published studies on the impact of this pandemic on anxiety levels in the UAE community. Furthermore, there is a dearth of global literature exploring the impact of such public health emergencies on the mental health of children. While severe COVID-19 illness in children is significant but far less frequent than in adults [12], still, their mental health may be disproportionately affected due to the changes in their daily routines, reduction in social contact and the anxiety associated with the disease. With the evidence of how parental anxiety can explain anxiety disorders in children and adolescents [13, 14]; and with the number of recent reports on the psychological impact of COVID-19 on adults, the mental well-being of children should not be ignored. Parent and teacher observations are important in screening for psychological and emotional disorders in children [15] and play a significant role in being key informants [16]. Furthermore, parents and teachers positively influence children’s well-being [17, 18].

In this study, we aim to explore anxiety levels among parents, teachers and the general community amid the COVID-19 pandemic in the UAE, as well as to identify emotional and anxiety disorders in children as reported by their parents and teachers. We will assess parents’ and teachers’ knowledge, beliefs and perceived risks related to COVID-19, health-protective and hygienic behaviors, perceived worries and fears as well as parental coping mechanisms used to reduce anxiety in children. This makes our study the first in the UAE and the region to discuss this aspect in the current pandemic.

## 2. Methods

### 2.1 Recruitment of participants

Convenience sampling was used to recruit participants into the study. Initially, we randomly selected and contacted 17 schools across the UAE, inviting them to participate in the study. However, with the sudden school closures and the transitioning to online learning, only 4 schools responded and participated in the study. Participants were invited to take part in an online survey via mass communications using email announcements through participating schools and universities and through social media posts (Facebook, Instagram and WhatsApp). Teachers, parents, and members of the general public throughout the UAE, aged 18 years and over, were invited to participate and were asked to pass on the survey link to their contacts. Data were collected from 24^th^ March to 15^th^ May 2020. The survey was administered via the survey monkey platform https://www.surveymonkey.com/r/9CB5GNQ [19], and each response came from a unique IP address to ensure that there were no duplicate entries. Through the survey link, the first page explained the research objectives and assured participants their responses were confidential. The minimum sample size needed for this cross-sectional study was 385, calculated for an expected prevalence of 50%, margin of error of 5%, and 95% confidence level.

### 2.2 Ethical approval and consent

The study was approved by the University of Sharjah Ethics Committee (approval number REC-20-03-12-01) and the United Arab Emirates University research ethics review board (ERS_2020_6098).

### 2.3 Data Collection

A structured questionnaire comprising 32 items was used in this study. The questions were divided into 8 domains; demographics, knowledge, beliefs and perceived risk related to COVID-19, health-protective and hygienic behaviors, precautionary measures, worry and fear associated with COVID-19, general health, validated self-reported anxiety screening scales (adults and children) and coping mechanisms. The questionnaire (Appendix 1) was translated into Arabic by a certified translator, and back translated to English to ensure accuracy of the translation. The final version of the questionnaire was piloted among 10 members of the general community to ensure clarity and consistency between survey items. To ensure face and content validity, the questionnaire was sent to a group of 10 experts which consisted of faculty, teachers, parents, and a mental health expert who reviewed the survey for content accuracy, length, clarity and comprehensiveness. Modifications were made to the phrasing of questions and response items based on recommendations from the expert panel for improving clarity and comprehensibility. The questionnaire took approximately 10 minutes to complete during the piloting phase.

#### Demographics

Information was collected on participants’ age, gender, educational level, emirate or country of residence, marital status, number of children and ages and levels of schooling for children. We also collected information on employment status, monthly income and health insurance. Participants were also requested to indicate if they were parents, a parent and teacher, teachers only or were neither parents nor teachers.

#### Knowledge, beliefs and perceived risk related to COVID-19

Participants were asked to answer “true”, “false”, or “don’t know” on questions related to COVID-19, such as, “*there is no specific treatment*” and “*I feel a sense of social responsibility by staying at home*”. Perceived risk from COVID-19 was assessed on a four-point Likert scale (very likely to not likely at all) where participants responded to the likelihood of contracting COVID-19, surviving COVID-19 or developing severe illness if infected.

#### Health-protective practices and hygienic behaviors

Participants were asked to describe how often they followed precautionary hygienic measures since the COVID-19 outbreak. Responses to seven questions (Covering mouth when sneezing/coughing, using hand sanitizer, washing hands with soap, wearing face masks, avoiding crowded areas and public transport and avoiding handshakes) were measured on a 4-point Likert scale ranging from always to never. These questions were modified versions of precautionary and hygienic behavior questions used in previous research during MERS-CoV, Swine Flu and SARS outbreaks [7, 9, 20, 21]. In order to categorize hygienic behavior into dichotomous categories a standard median split was carried out [22] with a median cut-off of 25. A value of ≥25 indicated high exhibiting behaviors in our study.

#### Worry and fear associated with COVID-19

To assess worry and fear associated with COVID-19, participants were asked to rate how worried they were on seven questions (*worried about catching COVID-19 myself, worried about parents catching COVID-19, worried about child catching COVID-19, worried about what COVID-19 can do to me health-wise, worried about social isolation/quarantine, worried about loss of income and worried about transmitting the virus to family and friends*). Participants rated their worry for each of these questions on a five-point Likert scale ranging from extremely worried to not worried at all). Additionally, participants were asked to describe (on a 5-point Likert scale, strongly agree to strongly disagree); their opinion on the public fear associated with the COVID-19 outbreak [23]. In order to categorize worry into dichotomous categories, a standard median split was carried out [22] with a median cut-off of 22. A value of 22 or above was used to signify very worried in our study.

#### Anxiety

Anxiety among adults was measured using the generalized anxiety disorder scale (GAD-7) [24] which is a self-reported 7-item validated scale. Participants were asked to indicate how often they were bothered during the previous 2 weeks by 7 symptoms of (*feeling nervous, not being able to stop worrying, worrying about different things, trouble relaxing, restless, irritable and afraid that something awful might happen*). Response options were “not at all,” “several days,” “more than half the days,” and “nearly every day,” scored as 0, 1, 2, and 3, respectively. A score of 10 or greater represents a reasonable cut point for identifying cases of anxiety with a sensitivity of 89% and specificity 82%, internal consistency (Cronbach α=.92) and Test-retest reliability (intraclass correlation=0.83) [24]. Other research has reported that a cutoff score of 8, (sensitivity 77% and specificity 82%) is a screener for any anxiety disorder including panic disorder, social anxiety phobia and post-traumatic stress disorder (PTSD) [25]. GAD-7 scores were totaled for each participant and classified into cut-off points of (0-4 minimal, 5-9 mild, 10-14 moderate and 15-21 severe) [24] levels of anxiety as well as stratified into 2 groups (below or above the cut-off point of 8).

Anxiety levels of children were measured using the emotional symptoms sub-scale from the strengths and difficulties questionnaire (SDQ) [26]. The SDQ is a brief screening questionnaire for mental health problems and covers a broad range of mental health symptoms including emotional symptoms, conduct problems, hyperactivity-inattention, peer relationship problems, and prosocial behaviors. It has been designed to screen for psychological disorders in children and is administered by parents and teachers of children aged 3 to 16 years [16]. The emotional symptoms sub-scale of the SDQ comprises of 5 questions which have been used to screen for anxiety disorders in children [27]. It asks parents and teachers questions about symptoms they have witnessed in children; (*Often complains of headaches, stomach-ache or sickness”; “Many worries, often seems worried”; “Often unhappy, down-hearted or tearful”; “Nervous or clingy in new situations, easily loses confidence”; and “Many fears, easily scared*). Each item can be marked “not true”, “somewhat true” or “certainly true” and is scored 0, 1 and 2 respectively thereby generating a scale score ranging from 0 to 10. A suggested cutoff score of 7 has been identified as a screener for generalized anxiety disorder (sensitivity 75% and specificity 80%) and depressive and generalized anxiety disorders (sensitivity 67% and specificity 81%) in children [28]. However, according to the tools scoring guidelines [29], an abnormal emotional problems score completed by both parents and teachers ranged from 5-10 and therefore we used an SDQ≥5 as an indicator of abnormal emotional score. Validated Arabic translations of both the GAD-7 and SDQ were used for the Arabic translation of the questionnaire [30, 31].

In order to determine the impact of precautionary measures undertaken by the government in reducing anxiety levels, participants were asked whether they felt less anxious with the introduction of (*online learning, airport screening, travel bans, availability of hand sanitizer in public places, cancellation of social events, temporary closure of public places and social isolation*). Responses were recorded on a 5-point Likert scale ranging from strongly agree to strongly disagree. In order to categorize precautionary measures into dichotomous categories, a standard median split was carried out [22] with a median cutoff of 34. A score of ≥34 indicated high agreement with precautionary measures in our study.

#### Coping mechanisms

Participants were asked to indicate on a 4-point Likert scale, (ranging from always to never) which coping mechanisms they had undertaken to help reduce anxiety in their children and family during the COVID-19 outbreak. Questions included, (*openly discussing COVID-19 with children/family, education children about proper hygienic measures, assuring children they are safe, limiting children’s exposure to news coverage including social media, creating a schedule of learning and fun activities and maintaining a regular routine at home)*. In order to categorize coping strategies into dichotomous categories a standard median split was carried out [22] with a median cutoff of 18. A score of ≥18 indicated high coping strategies.

#### General health

Participants were asked to indicate whether they suffered from any chronic diseases and to report any flu-like symptoms they had experienced over the previous 2 weeks, the measures taken for such symptoms (treatments), the likelihood of taking a COVID-19 vaccine if it was developed, whether their children were up to date with vaccinations and whether or not they smoked and if so, what they smoked. Participants were also asked to indicate if their smoking habits had changed since the COVID-19 outbreak.

### 2.4 Statistical analysis

Descriptive statistics, including means, medians, frequencies and percentages were used to summarize data and to illustrate the demographic and other selected characteristics of participants. The normal distribution of data was verified visually using histo-grams, boxplots, and quantile-quantile plots, statistically using the Kolmogorov-Smirnov test. and the equality of variances was checked using the Levene’s test. Bivariate analysis using Chi-square (χ2) tests explored the associations between participant demographic characteristics and anxiety levels. Statistically significant factors in the bivariate analysis were included in the multivariate binary logistic regression models to determine predictors of anxiety levels (GAD-7 score ≥8) and emotional problems in children (SDQ score ≥5). The automatic selection of predictors in the model was performed by a stepwise backward method with an entry threshold of 0.05 and an exit threshold of 0.1. The adequacy of the model was verified by the Hosmer and Lemeshow test and the specificity of the model by Link Test. The estimates of the strengths of associations were demonstrated by the odds ratio (OR) with a 95% confidence interval (CI). A two-tailed *p <0*.*05* was considered statistically significant. Data were analyzed using the statistical software SAS® 9.3 [32].

## 3. Results

In total, 2200 people completed the online participant sheet and consent form. Of these 26 declined to participate in the study and 381 completed only the demographic part of the questionnaire before discontinuing. Complete data were analyzed for a total of 1469 participants giving this research a completion rate of 68%.

A summary of the demographics for our sample is displayed in Table 1. Participants were primarily females (82.8%), predominantly from the 25 to 44 year age group (61.7%) and were residing in the United Arab Emirates (72.8%). Over half of our population held a bachelor’s degree (50.8%) and were employed (63.1%). Seventy five percent of participants were married and had children (75.6%), with the majority having one or two children (35.2%). The most commonly reported medical conditions were high blood pressure (9.1%) and asthma (8.6%). Headaches (25.9%) were the most commonly reported COVID-19 associated symptom and almost half of participants indicated they used Vitamin C for their symptoms. Whilst most participants reported they did not smoke, 13.7% stated they had changed their smoking habits since the COVID-19 outbreak. Most participants indicated they would vaccinate themselves (71.5%) and their children (59.4%) with the COVID-19 vaccine once developed. The majority of participants indicated their children were up to date with their vaccines (85%), however, we found a significant association between those who reported their children were not up-to-date with their vaccinations (53%) and their intention to not vaccinate their children with the COVID-19 vaccine once developed [χ^2^ (2, N=1469) = 31.05, p<0.001].

**Table 1.**
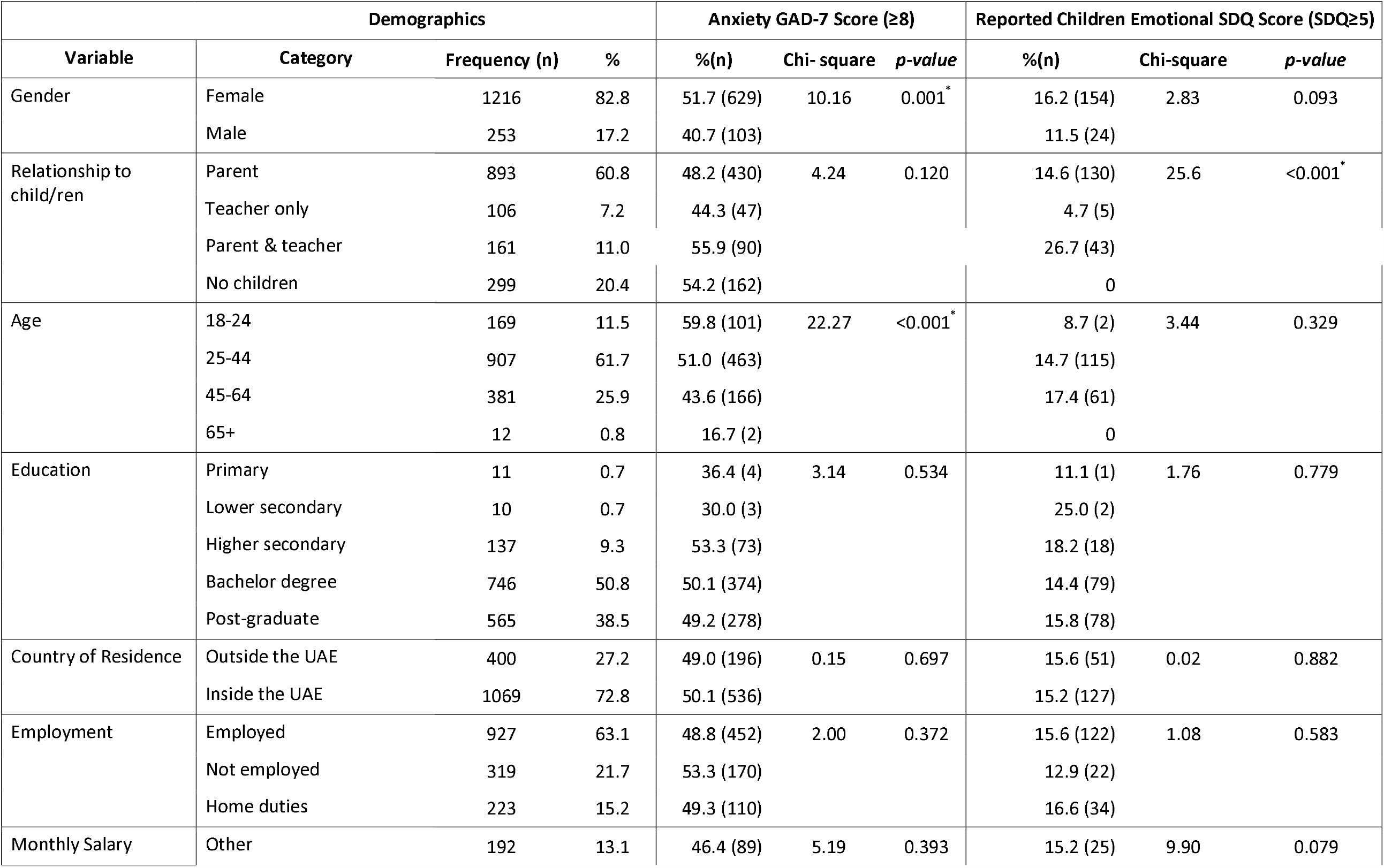
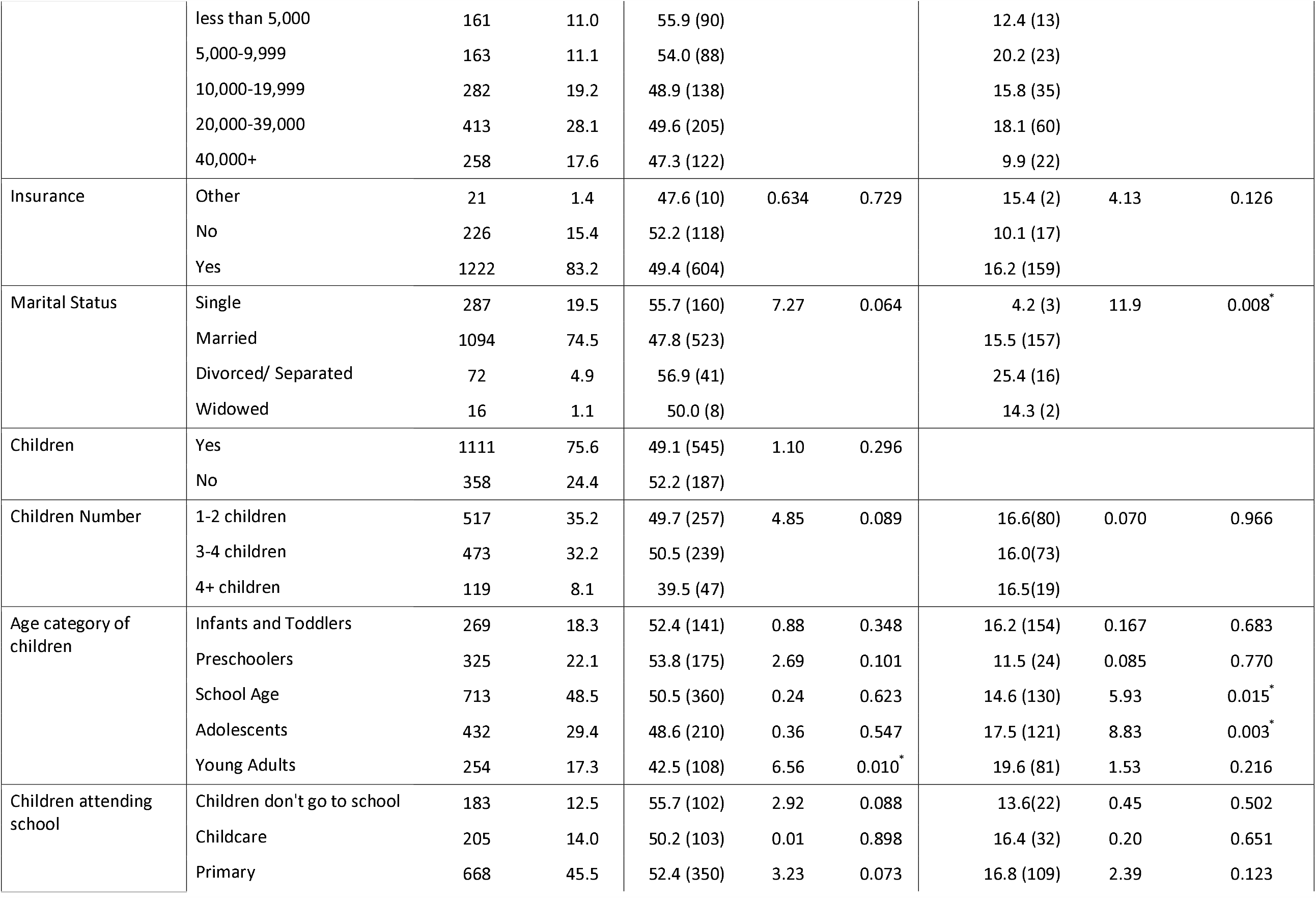
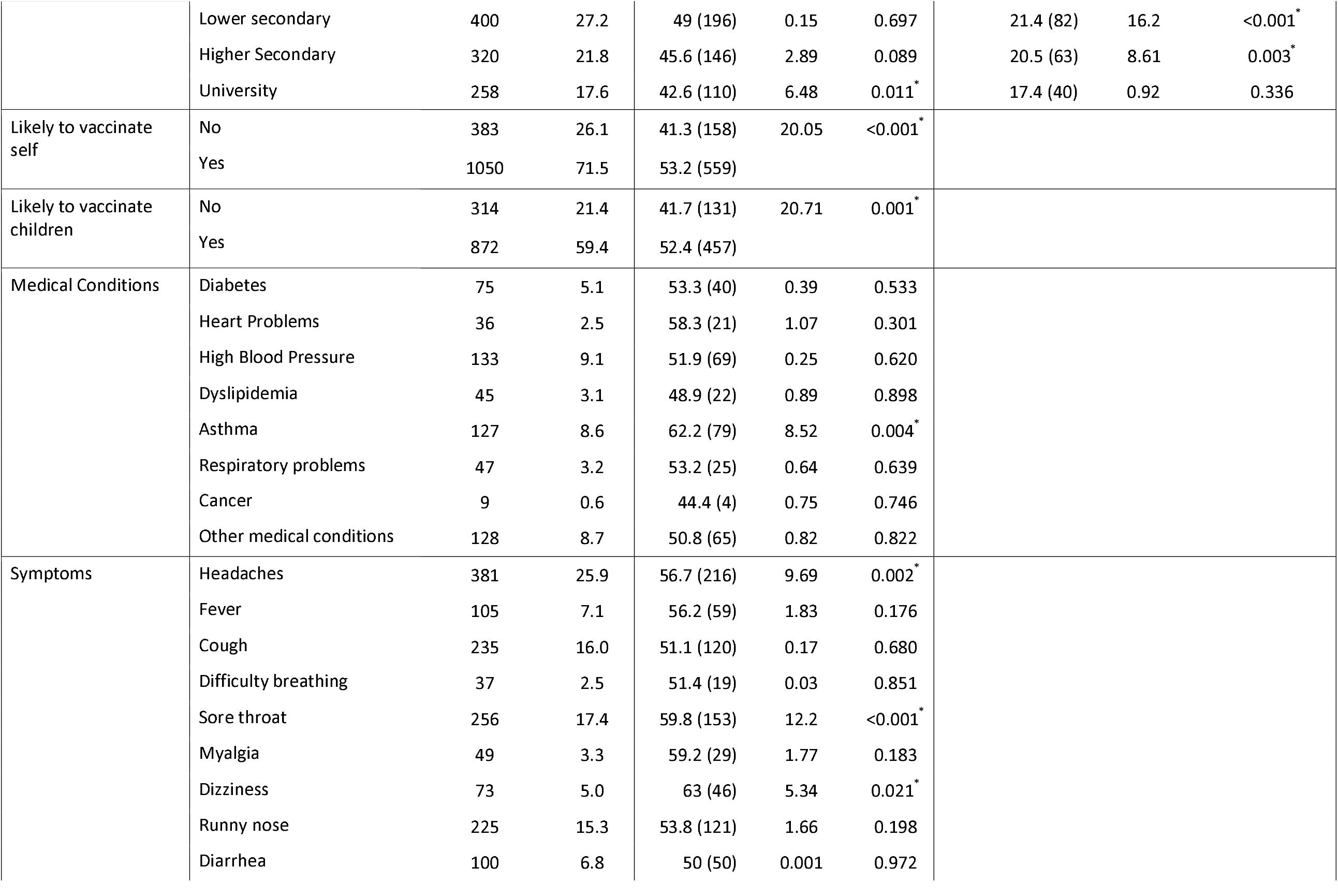
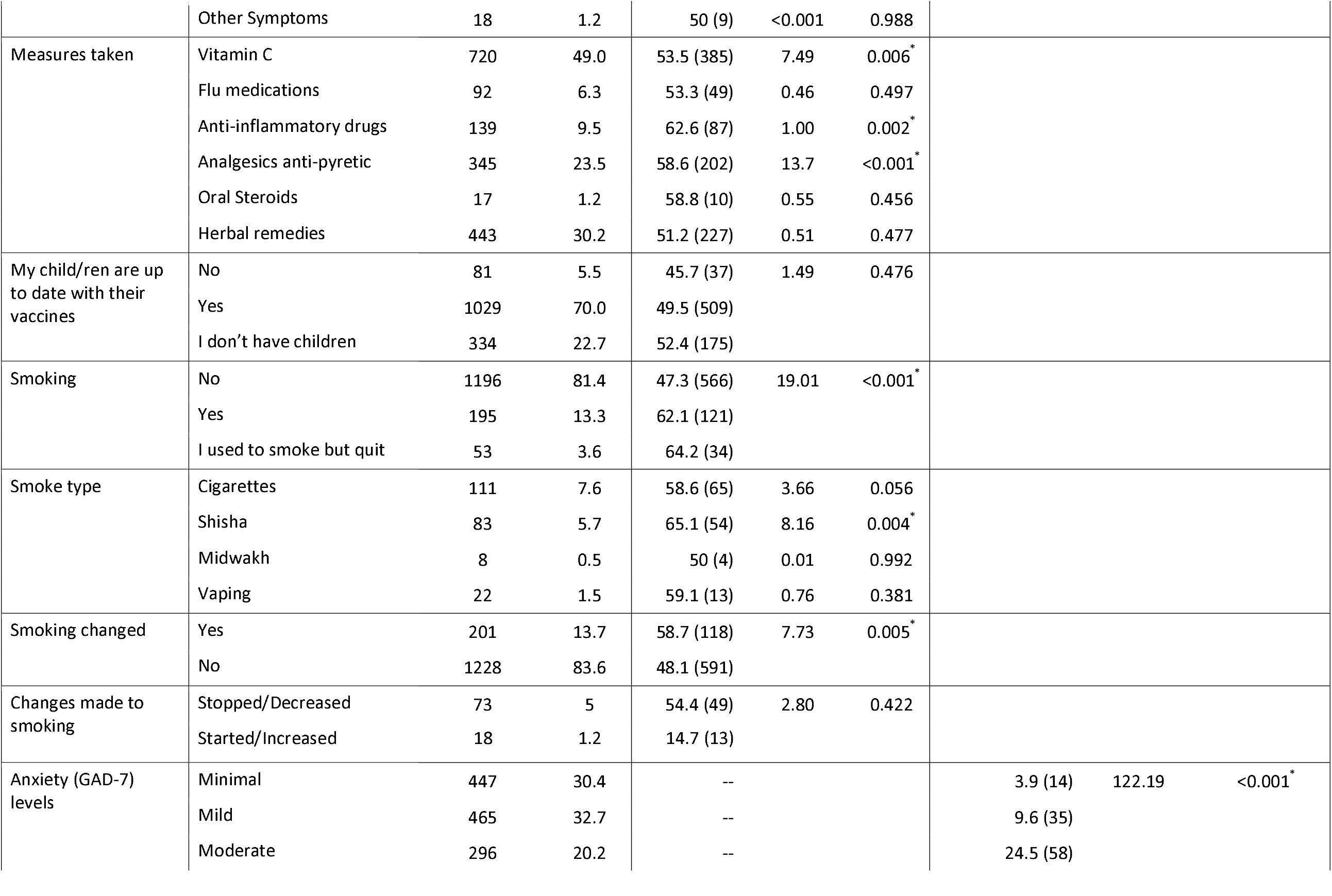
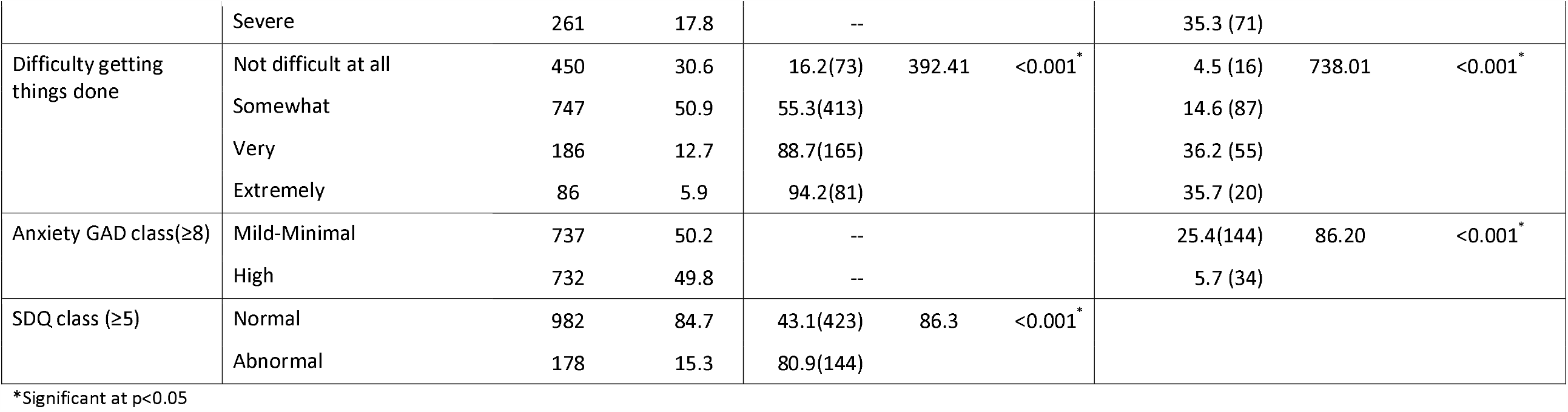
Demographic characteristics by anxiety score (GAD-7) and children emotional SDQ score *(n=1469)*

### Anxiety levels (GAD-7 score and SDQ score)

Almost three quarters (71%) of our population reported anxiety. Of these, thirty-eight percent reported moderate to severe anxiety. When we categorized anxiety by high and low based on the GAD-7 score cutoff point of a median of 8, half of our participants (49.8%) reported higher levels of anxiety. Females (51.7%) and participants between the ages of 18 and 24 years (59.8%) reported significantly higher levels of anxiety [χ^2^ (1, N=1469) = 10.16, p<0.001 and χ^2^ (5, N=1469) = 22.27, p<0.001 respectively]. Even though higher anxiety levels were reported amongst participants with higher levels of education, the differences were not significant [χ^2^ (4, N=1469) = 3.14, p =0.534]. More than half of participants who indicated they were likely to vaccinate themselves with the COVID-19 vaccine were more anxious [χ^2^ (2, N=1469) = 20.05, p ≤0.001]. Similarly, more than half of parents who indicated they were likely to vaccinate their children with the COVID-19 vaccine had higher anxiety levels, [χ ^2^ (1, N=1469) = 20.71, p<0.001]. Significantly higher levels of anxiety were reported in participants who had asthma [χ^2^ (1, N=1469) = 8.52, p=0.004], symptoms of headaches [χ^2^ (1, N=1469) = 9.69, p=0.002], sore throat [χ^2^ (1, N=1469) = 12.24, p≤0.001], and dizziness [χ^2^ (1, N=1469) = 5.34, p<0.001]. Participants who reported higher anxiety were more likely to take Vitamin C (53.5%), p=0.006), anti-inflammatory drugs (62.6%, p=0.002), and analgesics (58.6%, p<0.001) than those with lower levels of anxiety. Participants who indicated they used to smoke but had now quit, had higher anxiety levels [χ^2^ (2, N=1469)=19.01,p≤0.001]. Among those who smoked and reported higher anxiety levels, Shisha (waterpipe or Hookah smoking) was the most common type of smoking reported (65.1%), [χ^2^ (2, N=1469)=8.16, p=0.004] as displayed in Table 1.

The highest percentage of reported emotional problems for children using the SDQ score was in participants who were both parents and teachers (26.7%) compared to parents only(14.6%) or teachers only 4.7% [χ^2^ (2, N=1469) = 25.6, p<0.001]. Participants who were divorced/separated reported higher SDQ scores in their children (25.4%), compared to those who were married (15.5%) [χ^2^ (3, N=1469) = 11.88, p<0.008] and children who were school aged or adolescents showed significant differences in reports of emotional problems compared to children who were not school-aged or adolescents (17.5%) [χ^2^ (2, N=1469) = 5.93, p=0.015] and (19.6%) [χ^2^ (2, N=1469) = 8.83, p=0.003] respectively). Emotional problems were also more commonly reported in children attending lower secondary and higher secondary schools (p<0.001, p=0.003, respectively). Parents who reported moderate and severe anxiety levels in the GAD-7, also reported higher SDQ scores in their children χ^2^ (3, N=1469) = 122.19, p≤0.001). A higher percentage of parents who reported children with emotional problems also reported they found it “Very or extremely” difficult to get things done (36.1%) [χ^2^ (3, N=1469) = 101.22, p≤0.001] as displayed in Table 1.

### Knowledge, beliefs, hygienic behavior and anxiety levels

Overall, participants showed a good knowledge of COVID-19 and the majority were aware that there was no treatment. Participants (83%) perceived a likelihood of catching COVID-19 with almost half of these participants reporting higher levels of anxiety [χ^2^ (2, N=1469) =7.26, p=0.026]. More than half of participants who believed they would develop severe illness if they contracted the virus reported higher levels of anxiety [χ^2^ (2, N=1469) =13.56, p<0.001] (Table 2). Almost all participants had made significant changes in their hygienic behavior since the COVID-19 outbreak and reported increased use of hand sanitizer (87%), washing hands (99%), wearing face masks (47%), avoiding crowds (96%), public transport (98%) and handshaking (95%). Significantly higher levels of anxiety were reported amongst participants who reported always using hand sanitizers [χ^2^ (2, N=1469) = 10.90, p<0.001] and wearing face masks [χ^2^ (2, N=1469) = 8.84, p<0.001]. When behavioral changes were further categorized into 2 groups, participants who “always” practiced hygienic behaviors, reported significantly higher levels of anxiety [χ^2^ (1, N=1469) = 8.18, p<0.001] (Table 3).

**Table 2:**
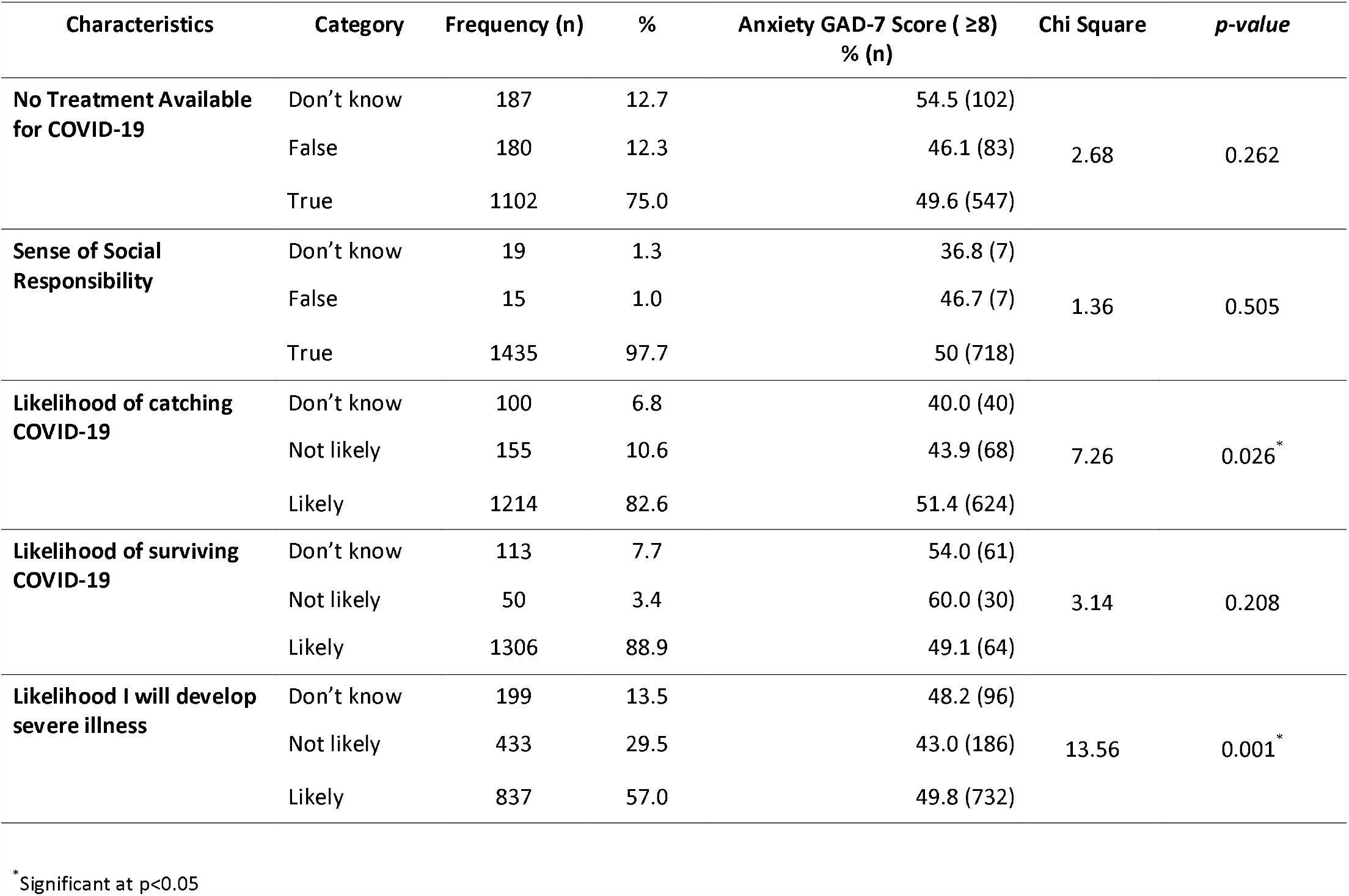
Prevalence of GAD-7 score ≥8 by knowledge and beliefs related to COVID-19 (n=1469)

| Characteristics | Category | Frequency (n) | % | Anxiety GAD-7 Score ($\geq 8$)<br>% (n) | Chi Square | *p-value* |
| --- | --- | --- | --- | --- | --- | --- |
| **No Treatment Available for COVID-19** | Don't know | 187 | 12.7 | 54.5 (102) | 2.68 | 0.262 |
|  | False | 180 | 12.3 | 46.1 (83) |  |  |
|  | True | 1102 | 75.0 | 49.6 (547) |  |  |
| **Sense of Social Responsibility** | Don't know | 19 | 1.3 | 36.8 (7) | 1.36 | 0.505 |
|  | False | 15 | 1.0 | 46.7 (7) |  |  |
|  | True | 1435 | 97.7 | 50 (718) |  |  |
| **Likelihood of catching COVID-19** | Don't know | 100 | 6.8 | 40.0 (40) | 7.26 | 0.026* |
|  | Not likely | 155 | 10.6 | 43.9 (68) |  |  |
|  | Likely | 1214 | 82.6 | 51.4 (624) |  |  |
| **Likelihood of surviving COVID-19** | Don't know | 113 | 7.7 | 54.0 (61) | 3.14 | 0.208 |
|  | Not likely | 50 | 3.4 | 60.0 (30) |  |  |
|  | Likely | 1306 | 88.9 | 49.1 (64) |  |  |
| **Likelihood I will develop severe illness** | Don't know | 199 | 13.5 | 48.2 (96) | 13.56 | 0.001* |
|  | Not likely | 433 | 29.5 | 43.0 (186) |  |  |
|  | Likely | 837 | 57.0 | 49.8 (732) |  |  |

\* Significant at $p < 0.05$

**Table 3.** Prevalence of GAD-7 score ≥8 by behavior changes taken (*n=1469*)

| Characteristics | Category | Frequency (n) | % | Anxiety GAD-7 Score ($\geq 8$)<br>% (n) | Chi-Square | *p-value*<sup>*</sup> |
| --- | --- | --- | --- | --- | --- | --- |
| **Cover mouth** | Never | 13 | 0.9 | 46.2 (6) | 3.03 | 0.219 |
|  | Occasionally | 43 | 2.9 | 62.8 (27) |  |  |
|  | Most of the time /Always | 1413 | 96.2 | 49.4 (699) |  |  |
| **Use hand sanitizer** | Never | 26 | 1.8 | 26.9 (7) | 10.90 | 0.004<sup>*</sup> |
|  | Occasionally | 163 | 11.1 | 41.7 (68) |  |  |
|  | Most of the time /Always | 1280 | 87.1 | 51.3 (657) |  |  |
| **Washing hands** | Never | 2 | 0.1 | 50.0 (1) | 1.13 | 0.569 |
|  | Occasionally | 14 | 1.0 | 35.7 (5) |  |  |
|  | Most of the time /Always | 1453 | 98.9 | 50.0 (726) |  |  |
| **Face mask** | Never | 374 | 25.5 | 48.9 (183) | 8.84 | 0.012<sup>*</sup> |
|  | Occasionally | 408 | 27.8 | 44.4 (181) |  |  |
|  | Most of the time /Always | 687 | 46.8 | 53.6 (368) |  |  |
| **Avoid crowds** | Never | 6 | 0.4 | 50.0 (3) | 0.25 | 0.882 |
|  | Occasionally | 60 | 4.1 | 46.7 (28) |  |  |
|  | Most of the time /Always | 1403 | 95.5 | 50 (701) |  |  |
| **Avoid public transport** | Never | 14 | 1.0 | 50.0 (7) | 0.17 | 0.918 |
|  | Occasionally | 22 | 1.5 | 45.5 (10) |  |  |
|  | Most of the time /Always | 1433 | 97.5 | 49.9 (715) |  |  |
| **Avoid handshaking** | Never | 12 | 0.8 | 66.7 (8) | 1.59 | 0.451 |
|  | Occasionally | 62 | 4.2 | 46.8 (29) |  |  |
|  | Most of the time /Always | 1395 | 94.9 | 49.8 (695) |  |  |
| **Behavioral changes total Category** | Occasionally exhibiting behavior changes | 604 | 41.1 | 45.4(274) | 8.18 | <0.004<sup>*</sup> |
|  | Always exhibiting behavior changes | 865 | 58.9 | 53.0(458) |  |  |

<sup>\*</sup>Significant at $p < 0.05$

### Perception of precautionary measures and anxiety levels

Although most participants in our study agreed that they felt less anxious with the precautionary measures introduced by the government, participants who disagreed reported higher GAD-7 scores. Particularly for online learning [χ^2^ (2, N=1469) = 36.55, p<0.001], cancellation of social events [χ^2^ (2, N=1469) = 7.59, p<0.001] and social isolation [χ^2^ (2, N=1469) = 7.26, p<0.001]. Participants who agreed overall with the precautionary measures introduced showed significantly lower levels of anxiety than those who disagreed [χ^2^ (1, N=1469) = 10.48, p<0.001] (Table 4).

**Table 4.** Prevalence of GAD-7 score ≥8 by precautionary measures taken *(n=1469)*

| Characteristics | Category | Frequency (n) | % | Anxiety GAD-7 Score ($\geq 8$)<br>% (n) | Chi Square | p-value |
| --- | --- | --- | --- | --- | --- | --- |
| Online learning at educational institutions | Strongly disagree/Disagree | 220 | 15.0 | 65.0 (143) | 36.55 | <0.001* |
|  | Neutral | 238 | 16.2 | 57.6 (137) |  |  |
|  | Strongly agree/Agree | 1011 | 68.8 | 44.7 (452) |  |  |
| Airport screening | Strongly disagree/Disagree | 67 | 4.6 | 61.2 (41) | 14.14 | 0.001* |
|  | Neutral | 136 | 9.3 | 62.5 (85) |  |  |
|  | Strongly agree/ Agree | 1266 | 86.2 | 47.9 (606) |  |  |
| Travel bans | Strongly disagree/Disagree | 65 | 4.4 | 55.4 (36) | 11.11 | 0.004* |
|  | Neutral | 79 | 5.4 | 67.1 (53) |  |  |
|  | Strongly agree/Agree | 1325 | 90.2 | 48.5 (643) |  |  |
| Hand sanitizers in public spaces | Strongly disagree/ Disagree | 33 | 2.2 | 51.5 (17) | 2.31 | 0.315 |
|  | Neutral | 98 | 6.7 | 57.1 (56) |  |  |
|  | Strongly agree/ Agree | 1338 | 91.1 | 49.3 (659) |  |  |
| Cancellation of social events | Strongly disagree/Disagree | 34 | 2.3 | 61.8 (21) | 7.59 | 0.022* |
|  | Neutral | 50 | 3.4 | 66.0 (33) |  |  |
|  | Strongly agree /Agree | 1385 | 94.3 | 49.0 (678) |  |  |
| Temporary closure of public places | Strongly disagree/Disagree | 47 | 3.2 | 51.1 (24) | 1.76 | 0.415 |
|  | Neutral | 51 | 3.5 | 58.8 (30) |  |  |
|  | Strongly agree/Agree | 1371 | 93.3 | 49.5 (678) |  |  |
| Social isolation | Strongly disagree /Disagree | 49 | 3.3 | 61.2 (30) | 7.26 | 0.026* |
|  | Neutral | 82 | 5.6 | 61.0 (50) |  |  |
|  | Strongly agree/Agree | 1338 | 91.1 | 48.7 (652) |  |  |
| Precautionary measures category | Disagree with precautionary measures | 605 | 41.2 | 54.9(332) | 10.48 | 0.001* |
|  | Agree with precautionary measures | 864 | 58.8 | 46.3(400) |  |  |

\* Significant at $p < 0.05$

### Worry and fear and anxiety levels

Overall, public fear was justified among participants in our study with the majority agreeing that the public fear was appropriate. However, we found higher levels of anxiety among those who believed that the fear had caused unnecessary absences from work and schools [χ^2^ (2, N=1469) = 10.86, p<0.001]. Whilst most participants reported being worried about catching COVID-19, the majority were more worried about their parents (75%) or children (65.5%) catching COVID-19 or transmitting it to someone else if they caught it (64.5%). Significantly higher GAD-7 scores were found among all participants who agreed they were worried about catching COVID-19, their parents or children catching it, worried about what it would do to them if they caught it, worried about being in social isolation, loss of income and transmitting it to others. When we categorized worry into 2 groups, “not too worried” and “very worried”, we found significantly higher levels of anxiety among participants who reported being very worried [χ^2^ (1, N=1469) = 148.7, p≤0.001] (Table 5). Worry in parents was also found to be associated with the SDQ score. Parents who reported higher levels of worry also reported higher emotional problems in their children on all the worry questions. In the worry category, parents who were very worried reported significantly higher SDQ scores for their children [χ^2^ (1, N=1469) = 44.90, p≤0.001] (Table 5).

**Table 5.** Worry about COVID-19 by GAD-7 score and SDQ score *(n=1469)*

| Characteristics | Category | Frequency (n) | % | Anxiety GAD-7 Score (≥8) |  |  | Reported Children Emotional SDQ Score (SDQ≥5) |  |  |
| --- | --- | --- | --- | --- | --- | --- | --- | --- | --- |
|  |  |  |  | %(n) | Chi-Square | p-value | %(n) | Chi-square | p-value |
| Public fear is justifiable | Strongly disagree/Disagree | 85 | 5.8 | 31.8 (27) | 29.08 | <0.001* | 29.7 (11) | 0.89 | 0.640 |
|  | Neutral | 156 | 10.6 | 35.3 (55) |  |  | 9.5(71) |  |  |
|  | Strongly agree /Agree | 1228 | 83.6 | 52.9 (650) |  |  | 29.2 (186) |  |  |
| Public fear is dysfunctional | Strongly disagree/Disagree | 759 | 51.7 | 51.5 (391) | 10.86 | 0.004* | 28.5 (107) | 0.357 | 0.836 |
|  | Neutral | 261 | 17.8 | 40.6 (106) |  |  | 13.9(29) |  |  |
|  | Strongly agree/ Agree | 448 | 30.6 | 52.3 (235) |  |  | 30 (74) |  |  |
| Worried catching COVID-19 | Not worried at all | 138 | 9.5 | 21.7 (30) | 176.98 | <0.001* | 6.6 (7) | 55.25 | ≤0.001* |
|  | Little/Somewhat worried | 801 | 55.2 | 40.6 (325) |  |  | 10.3 (67) |  |  |
|  | Very/Extremely worried | 511 | 35.2 | 72.6 (371) |  |  | 26.3 (104) |  |  |
| Worried parents catch COVID-19 | Not worried at all | 49 | 3.6 | 14.3 (7) | 98.86 | ≤0.001* | 5.1 (2) | 18.24 | ≤0.001* |
|  | Little/Somewhat worried | 294 | 21.4 | 29.6 (87) |  |  | 8.2(20) |  |  |
|  | Very/Extremely worried | 1028 | 75.0 | 57.8(594) |  |  | 18.4 (146) |  |  |
| Worried children catch COVID-19 | Not worried at all | 62 | 5.3 | 19.4 (12) | 110.44 | ≤0.001* | 4 (2) | 23.14 | ≤0.001* |
|  | Little/Somewhat worried | 342 | 29.2 | 29.8(102) |  |  | 10 (32) |  |  |
|  | Very/Extremely worried | 766 | 65.5 | 60.2(461) |  |  | 20.4 (141) |  |  |
| Worried what COVID-19 can do to me healthwise | Not worried at all | 121 | 8.4 | 17.4 (21) | 178.81 | ≤0.001* | 3.1 (3) | 44.22 | ≤0.001* |
|  | Little/Somewhat worried | 667 | 46.4 | 38.4 (256) |  |  | 10.5 (56) |  |  |
|  | Very/Extremely worried | 647 | 45.1 | 68.8 (445) |  |  | 23.1 (117) |  |  |
| Worried about social isolation | Not worried at all | 375 | 26.1 | 34.4(129) | 81.71 | ≤0.001* | 9.2 (27) | 58.29 | ≤0.001* |
|  | Little/Somewhat worried | 608 | 42.3 | 48.2(293) |  |  | 10.6 (52) |  |  |
|  | Very/Extremely worried | 454 | 31.6 | 65.6(298) |  |  | 27.7 (99) |  |  |
| Worried about loss of income if infected with COVID-19 | Not worried at all | 265 | 18.8 | 32.1 (85) | 73.63 | ≤0.001* | 7.9 (16) | 22.91 | ≤0.001* |
|  | Little/Somewhat worried | 441 | 32.9 | 44.4 (196) |  |  | 12.1 (44) |  |  |
|  | Very/Extremely worried | 633 | 47.3 | 61.6 (390) |  |  | 20.7(107) |  |  |
| Worried I transmit COVID-19 | Not worried at all | 104 | 8.5 | 31.7 (33) | 79.81 | ≤0.001* | 8.5 (8) | 32.48 | ≤0.001* |
|  | Little/Somewhat worried | 328 | 27.0 | 34.5 (113) |  |  | 7.4 (20) |  |  |
|  | Very/Extremely worried | 785 | 64.5 | 60.5 (475) |  |  | 21.6 (131) |  |  |
| Worried in total about COVID-19 | Low levels of worry | 394 | 26.8 | 27.4 (108) | 148.7 | $\leq 0.001^*$ | 6.4 (18) | 44.9 | $\leq 0.001^*$ |
|  | High levels of worry | 1075 | 73.2 | 58 (624) |  |  | 18.2 (160) |  |  |

\* Significant at $p < 0.05$

We found that among participants who had children, most were utilizing effective coping strategies. However; significantly higher anxiety was reported among participants who always openly discussed COVID-19 with their family (51.4%), compared to those who never did (33.3%, *p=0*.*013*). Similarly we found higher anxiety levels among participants who indicated they always educated their children about proper protective measures (50.3%), or limited their children’s news exposure (53.4%) compared to those who never did any of these (23.1%, *p=0*.*017*) and (41%, p=0.011) respectively. When we categorized all these strategies into 2 groups “occasionally” and “always”, we found no differences in anxiety levels based on the GAD-7 score among participants [χ^2^ (1, N=1469) = 1.65, p=0.199]. For the SDQ scores reported by parents, we found that higher emotional problems were reported in children whose parents/teachers discussed COVID-19 with them and among those who educated their children about personal protective measures (17.5%, p=0.009, 20.9%, p<0.001, respectively). Overall, parents who always utilized coping strategies for dealing with information regarding COVID-19 reported children with higher emotional problems as compared to parents who utilized fewer coping strategies [χ^2^ (1, N=1469) = 9.01, p≤0.001] (Table 6).

**Table 6.**
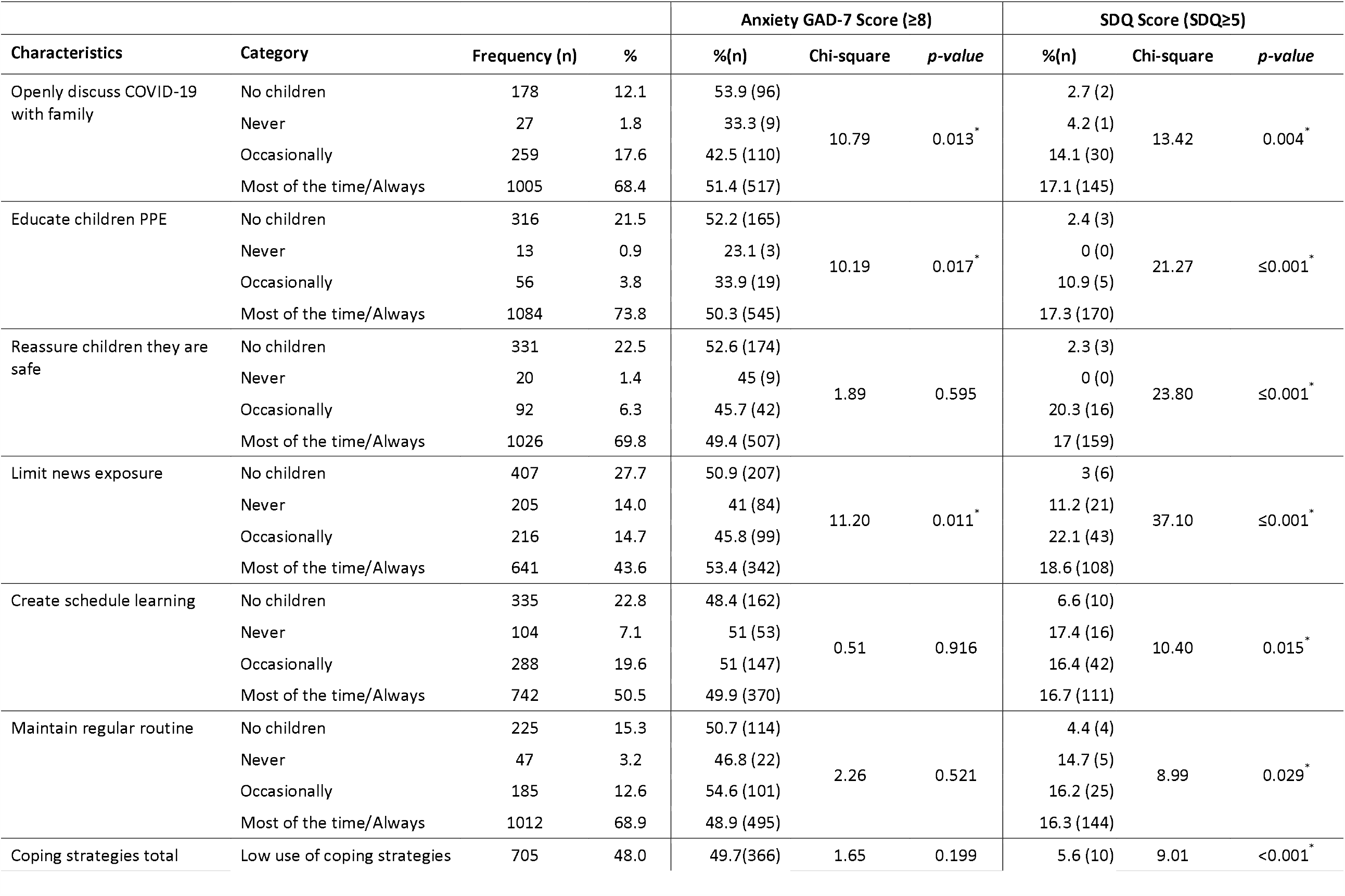
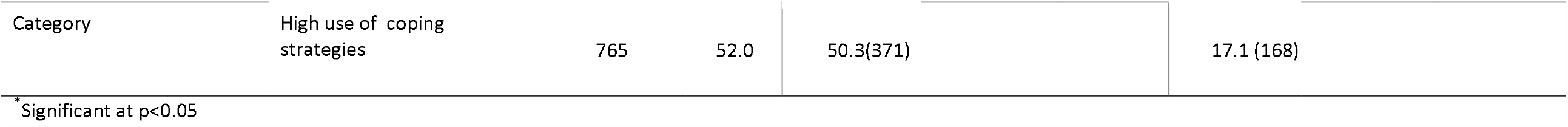
Coping strategies used with children by GAD-7 score and SDQ score (n=1469)

In order to estimate the probability of anxiety levels among participants in our study, 2 multivariate logistic regressions were conducted. One with the (GAD-7 ≥ 8) score as a measure of anxiety in adults and the other with the (SDQ ≥ 5) score as a measure of anxiety and emotional problems in children. For the first model, the effects of gender, age, age of children, perception of fear, perception of likelihood to contract COVID-19 and to develop severe disease, headaches, sore throat, asthma, measures taken for symptoms, smoking, and changed smoking habits, likelihood of vaccination for self and children, hygienic behavior category, precautionary measures category and worry category were modelled. The omnibus model for the logistic regression analysis was statistically significant, [χ^2^ (40, N=1469) =276.2, p≤0.001]. The model explained 28% (Nagelkerke R^2^) of the variance in anxiety levels. Hosmer and Lemeshow test results confirmed that the model was a good fit for the data, χ^2^(8, N=1469) =7.16, *p*=0.519. Coefficients for the model’s predictors are presented in (Table 7). Females had 1.91 times higher odds of reporting anxiety than males and, participants who believed that the public fear was justifiable, were 6 times more anxious than those who disagreed. Higher levels of worry were also associated with increased anxiety levels (OR=3.80, 95% CI, 2.90 to 5.00). Participants who reported they would take the COVID-19 vaccine were 1.57 times more likely to report higher anxiety, however, vaccinating children was not found to influence anxiety levels (p=0.158). The odds of reporting higher anxiety were more likely among participants who smoked (OR=1.55, 95% CI, 1.06 to 2.26), were taking vitamin C for flu-like symptoms (OR=1.41, 95% CI, 1.09 to 1.83) and reported a sore throat (OR=1.56, 95% CI, 1.17 to 2.09) (Table 7).

**Table 7.** Multivariate analysis for variables predicting anxiety (GAD-7 score≥8) in sample population *(n=1469)*

| Variable |  | *b* | *SE(b)* | *P-value* | *OR [95% CI]* |
| --- | --- | --- | --- | --- | --- |
| **Gender** | Female | 0.649 | 0.178 | <0.001 | 1.91 [1.35-2.71] |
|  | Male<sup>a</sup> | -- | -- | -- | 1 |
| **Precautionary Measures** | Agree | -0.740 | 0.146 | <0.001 | 0.48 [0.36-0.63] |
|  | Disagree<sup>a</sup> | -- | -- | -- | 1 |
| **Public fear Justifiable** | Agree | 1.811 | 1.082 | 0.094 | 6.11 [0.73-51.0] |
|  | Disagree<sup>a</sup> | -- | -- | -- | 1 |
| **Worry associated with COVID-19** | High levels of worry | 1.336 | 0.139 | <0.001 | 3.80 [2.90-5.00] |
|  | Low levels of worry<sup>a</sup> | -- | -- | -- | 1 |
| **Will take COVID-19 Vaccine** | Yes | 0.446 | 0.1478 | 0.003 | 1.57 [1.17-2.09] |
|  | No<sup>a</sup> | -- | -- | -- | 1 |
| **Symptoms-Sore throat** | Yes | 0.447 | 0.173 | 0.010 | 1.56 [1.17-2.09] |
|  | No<sup>a</sup> | -- | -- | -- | 1 |
| **Taking Vitamin C as measure** | Yes | 0.344 | 0.134 | 0.010 | 1.41 [1.09-1.83] |
|  | No<sup>a</sup> | -- | -- | -- | 1 |
| **Smoker** | Yes | 0.435 | 0.194 | 0.025 | 1.55 [1.06-2.26] |
|  | No<sup>a</sup> | -- | -- | -- | 1 |
| Likelihood Ratio Chi-Square 276.2 (df=40, p-value≤0.001) |  |  |  |  |  |

<sup>a</sup> reference group, *b* parameter estimate, *SE*
Std Error, *OR* Odds Ratio, *CI* Confidence Interval

In the second model, with SDG≥5 as a measure of anxiety in children, the effects of relationship with children, age of children (school aged and adolescent aged), marital status, educational level of child (lower secondary and higher secondary), coping strategies, worry, parental GAD-7 anxiety level, and parental difficulty getting things done were modelled. The omnibus model for the logistic regression analysis was statistically significant, χ^2^ (17, N=1469) =185.90, p≤0.001. The model explained 26% (Nagelkerke R^2^) of the variance in parent reported anxiety levels in children. Hosmer and Lemeshow test results confirmed that the model was a good fit for the data, χ^2^ (7, N=1469) =11.99, *p*=0.101. Coefficients for the model’s significant predictors are presented in (Table 8). Adults who were both parents and teachers were five times more likely to report emotional problems in children (OR=5.08, 95% CI, 1.84 to 14.0) and the odds of reporting emotional problems were more likely in adolescents who were in lower and secondary education. Parents who reported anxiety levels in the GAD-7 were more likely to report higher emotional problems in their children. Parents who had severe anxiety levels were 7 times more likely to report more emotional problems in their children (OR=7.00, 95% CI, 3.45 to 14.0). Parental report of “finding it very difficult to do work, to do things at home and to get along with other people” was a strong predictor of emotional problems in children (OR=4.07, 95% CI, 2.10-8.05) (Table 8).

**Table 8.** Multivariate analysis for variables predicting parent/teacher reported emotional problems in children (SDQ score) *(n=1469)*

| Variable |  | *b* | *SE(b)* | *P-value* | *OR [95% CI]* |
| --- | --- | --- | --- | --- | --- |
| Relationship to child/ren | Parent only | 0.854 | 0.493 | 0.884 | 2.35 [0.89-6.17] |
|  | Parent & Teacher | 1.626 | 0.519 | <0.001 | 5.08 [1.84-14.0] |
|  | Teacher only<sup>a</sup> | -- | -- | -- | 1 |
| Child/ren in lower secondary education | Yes | 0.522 | 0.189 | 0.006 | 1.69 [1.16-2.44] |
|  | No<sup>a</sup> | -- | -- | -- | 1 |
| Child/ren in higher secondary education | Yes | 0.460 | 0.199 | 0.021 | 1.59 [1.07-2.34] |
|  | No<sup>a</sup> | -- | -- | -- | 1 |
| Anxiety level (GAD-7) | Severe | 1.94 | 0.355 | <0.001 | 7.00 [3.45-14.0] |
|  | Moderate | 1.505 | 0.340 | 0.013 | 4.51 [2.31-8.80] |
|  | Mild | 0.582 | 0.344 | 0.011 | 1.79 [0.91-3.50] |
|  | Minimal<sup>a</sup> | -- | -- | -- | 1 |
| Difficulty of parent/teacher to get things done | Extremely | 1.299 | 0.439 | 0.003 | 3.70 [1.55-8.66] |
|  | Very | 1.403 | 0.348 | <0.001 | 4.07 [2.10-8.05] |
|  | Somewhat | 0.805 | 0.306 | 0.009 | 2.24 [1.23-4.08] |
|  | Not difficult at all<sup>a</sup> | -- | -- | -- | 1 |
| Likelihood Ratio Chi-Square 185.90 (df=17, p-value≤0.001) |  |  |  |  |  |

<sup>a</sup> reference group, *b* parameter estimate, *SE* Std Error, *OR* Odds Ratio, *CI* Confidence Interval

## 4. Discussion

This study has revealed that the COVID-19 pandemic has made a significant impact on the mental health and well-being of the UAE population with the majority of participants in our study reporting mild to severe anxiety and scoring above the cut-off point of 8 on the GAD-7 scale. This finding was most prevalent among women in our study with reported higher levels of anxiety suggesting that females are suffering greater psychological impact from this pandemic than males which is consistent with reports from previous studies [33, 34]. In light of the current pandemic, increased anxiety in females may be further exacerbated by added pressures imposed on them to adapt to lifestyle changes in the context of COVID-19, with the added responsibility of online learning and home schooling as well as possibly managing work commitments, social isolation and increased fear and uncertainty for family and loved ones.

Overall, our study found that precautionary measures implemented by the government served as a protective factor for anxiety with most participants agreeing that they felt less anxious with the enforcement of these measures. However, those who disagreed that they felt less anxious, reported higher levels of anxiety associated with the introduction of online learning, airport screening and travel bans This may be due to the potential ramifications experienced in a community with the introduction of these measures, particularly over concerns with disrupted children’s education, online learning and the uncertainty of school examinations. Furthermore, with the UAE being a well-known and popular travel hub, and home to over 7 million expatriates, the airport closures and travel bans might have triggered increased anxiety in our participants since travel restrictions cause financial challenges and uncertainty with potential loss of jobs, suspended travel to be with family and loved ones and overall perception of lack of connectivity with the rest of the world causing concern and worry.

Worry is a mental process that has been reported as a key indicator of anxiety and a central feature of generalized anxiety disorder (GAD) [35-37]. In our study, high level of worry was a strong predictor for higher GAD-7 scores with participants reporting significant concerns for their parents and children contracting COVID-19, fears of transmitting COVID-19 from their workplace to their homes as well as concerns over the loss of income if infected with COVID-19. Worry in parents was associated with higher reported emotional problems in children, however, these findings faded when further analysis was done.

Higher risk perception was also found to be associated with high levels of anxiety in our study. Participants perceived a high risk of being infected with COVID-19 and if infected, they perceived a high risk of developing severe disease. These findings contradict previous research conducted in China during the early stages of the pandemic where participants reported lower perceived likelihood of contracting COVID-19 which was associated with lower levels of stress [11]. The high-risk perception among participants in our study could also explain the high compliance and uptake of protective and hygienic behaviors. Earlier research indicates that people who were more anxious about COVID-19 were also more engaged with regular hand hygiene and social distancing behaviors [38]. In the present study, more than half of participants who had higher GAD-7 scores, reported wearing face masks and regularly using hand sanitizers. The under or overestimation of the health risks associated with COVID-19 may reduce or increase the likelihood of complying with such measures on an individual basis often related to individual perceived health risks [39]. Those with pre-existing health conditions are more likely to perceive a higher sense of panic and concern due to perceived personal probability of infection [40]. This was demonstrated in our study where participants who reported asthma also reported higher anxiety revealing an awareness of risk perception and the probable development of emotional distress. Similarly, participants who reported headaches, sore throat, dizziness, taking vitamin C, smoking or recently quit smoking were more anxious than those who did not report these symptoms or practices. Sore throat, taking vitamin C and smoking remained significant predictors of anxiety levels among participants in our study when further analysis was conducted. Smoking, has been associated with adverse COVID-19 disease prognosis and smokers are at a higher risk of developing severe COVID-19 complications [41, 42]. The COVID-19 pandemic is a perfect opportunity for public health interventions and health care professionals to play a crucial role in addressing smoking cessation programs where healthy behavior is more likely to change positively due to affective measures of risk which predict protective behavior uptake [23]. A recent google trends study identified increased number of searches relating to smoking cessation showing an increased interest to quit smoking since the COVID-19 pandemic [43].

With the uncertainty of the pandemic and the race against time around the world to develop a vaccine for COVID-19, we found that the majority of our study population had the intention to vaccinate themselves and their children once the COVID-19 vaccine was available. Participants who reported higher anxiety were more likely to vaccinate although a relatively large percentage of participants also reported they had no intention to take the vaccine. This is similar to findings in a recent study in France that showed almost one quarter of the French population did not intend to use the COVID-19 vaccine when available [44] and findings from a local study showing a 12% vaccine hesitancy among the UAE population [45]. The majority of participants who had no intention to take the COVID-19 vaccine in our study also reported their children were not up to date with their vaccines. In order for a vaccine to successfully achieve the goal of population immunity, large-scale uptake in a community is crucial. In this study we did not explore the reasons behind the choices made by participants regarding their intentions to vaccinate or not, however recent studies have shown that hesitancies associated with a potential COVID-19 vaccine were mainly related to safety and political concerns [44, 46] as well as the already existing anti-vaccination movements which have strong pre-COVID-19 foundations. To overcome these potential fears and to ensure adequate uptake of the vaccine; it is essential that governments communicate the measures and processes to be taken to ensure the safety of the COVID-19 vaccine openly and transparently and implement strategies that increase vaccine confidence among the population.

For the first time, since the onset of this pandemic, the psychological impact of COVID-19 on children was assessed in our study. For the first time, our study provides information highlighting that COVID-19 has several adverse impacts on the mental health of children, particularly children in their lower and secondary levels of education who demonstrated higher emotional and psychological impact of COVID-19. This age group comprises of students most probably affected by social isolation, prolonged school closure, possible challenges with online learning and uncertainty or difficulty with assessments and examinations. Previous research has already revealed higher incidence of anxiety-related disorders among school-aged adolescents in the UAE [47] which makes this group especially vulnerable to the psychological impact of COVID-19. Although we did not interview children directly, we used validated parental/teacher questionnaires which have been validated against structured diagnostic interviews. Parents and teachers and especially parents who are teachers, were the best informants for screening emotional problems in children in our study. This is consistent with the literature suggesting that parents and teachers are key informants for reporting and screening potential psychological and emotional disorders in children [15, 16] and should be further trained in providing children with strategies for addressing these disorders. We found that parents who were regularly utilizing coping strategies with their children reported higher SDQ scores than those who were less likely to use these strategies. It is uncertain whether this demonstrates that these parents are more attuned with their children’s feelings because they are addressing these concerns therefore possibly over-reporting emotional symptoms or whether the strategies being used are not effective in reducing anxiety disorders in children. Further research should measure the effectiveness of these strategies in addressing anxiety disorders in children.

Furthermore, we found that higher parental anxiety was a significant predictor of children’s SDQ score suggesting a significant association between parental and child anxiety. This finding is consistent with previous research where mental health service utilization among adolescents was associated with parental anxiety and depression [13]. Our study also demonstrates that parental anxiety might be a unique factor and predictor in explaining anxiety disorders in children and adolescents and should be considered in future interventions and research investigating the psychological impact of public health emergencies in this population.

### Limitations

Despite the findings of this study, we acknowledge that it has several limitations. Firstly, the use of convenience sampling and its descriptive nature through an online survey may not allow the generalization of results. However, considering the need for a rapid method to assess the psychological impact on a population during a rapidly evolving infectious disease outbreak, the use of an online survey serves as a promising method for quick results [48]. Additionally, responses were collected from all over the UAE in addition to countries outside the UAE with good response rate allowing for a certain element of representation. Secondly, the nature of self-reported data in the survey may lead to response biases specifically for reported behavioral changes, coping strategies and measures taken where participants may provide socially desirable results. Furthermore, self-reported levels of anxiety and worry among adults and the perceived reports of emotional disorders for children, may not be as accurate as being assessed and evaluated by specialized health professionals. However, even with these limitations, this study provides important baseline information which will inform further research and public health interventions in this area.

## Conclusion

This is the first study to provide information on the initial response and psychological impact of the COVID-19 pandemic among adults and children, with significant association between parental and child anxiety. Worry and perception of public fear were found to be significant predictors of anxiety levels on our population. The findings can be used by policy makers to develop effective screening and coping strategies and to formulate interventions that improve the mental health of the population and specifically prevention programs for vulnerable children. Such strategies may reduce the psychological impact during the COVID-19 pandemic and other public health emergencies in the future.

## Data Availability

Data will be available upon request

## Author Contributions

BS, RH, FS, and MT conceived, designed and initiated the study. AA, IE, AH, ES contributed to the planning and implementation of the study. AAS analyzed survey data. BS, AAS interpreted the results. BS drafted the manuscript with input from RH, IE, AA, AH, MT, ES and QH. All authors read and approved the final version of the manuscript.

## Acknowledgements

The authors would like to thank all participants in the study.

## Supplementary Material

The survey used to collect data for this study is provided as supplementary material.

## References

1. WHO. World Health Organization. Coronavirus disease (COVID-19) outbreak. https://covid19whoint/.3-5-2020.

2. WHO. World Health Organisation Director General’s opening remarks at the media briefing on COVID-19, 11th March 2020. https://wwwwhoint/dg/speeches/detail/who-director-general-s-opening-remarks-at-the-media-briefing-on-covid-1911-march-2020.30-3-2020.

3. Worldometers.info. COVID-19 CORONAVIRUS PANDEMIC. Dover, Delaware, U.S.A. https://wwwworldometersinfo/coronavirus/.31-5-2020.

4. Wang C, Pan R, Wan X, Tan Y, Xu L, McIntyre RS, et al. A longitudinal study on the mental health of general population during the COVID-19 epidemic in China. Brain, behavior, and immunity. 2020. Epub 2020/04/17. doi:10.1016/j.bbi.2020.04.028. PubMed PMID: 32298802; PubMed Central PMCID: PMCPMC7153528.

5. Rubin GJ, Wessely S. The psychological effects of quarantining a city. BMJ (Clinical research ed). 2020;368:m313. Epub 2020/01/30. doi:10.1136/bmj.m313. PubMed PMID: 31992552.

6. Sim K, Huak Chan Y, Chong PN, Chua HC, Wen Soon S. Psychosocial and coping responses within the community health care setting towards a national outbreak of an infectious disease. Journal of psychosomatic research. 2010;68(2):195–202. Epub 2010/01/29. doi:10.1016/j.jpsychores.2009.04.004. PubMed PMID: 20105703; PubMed Central PMCID: PMCPMC7094450.

7. Balkhy HH, Abolfotouh MA, Al-Hathlool RH, Al-Jumah MA. Awareness, attitudes, and practices related to the swine influenza pandemic among the Saudi public. BMC infectious diseases. 2010;10:42. Epub 2010/03/02. doi:10.1186/1471-2334-10-42. PubMed PMID: 20187976; PubMed Central PMCID: PMCPMC2844401.

8. Lau JT, Yang X, Tsui HY, Pang E. SARS related preventive and risk behaviours practised by Hong Kong-mainland China cross border travellers during the outbreak of the SARS epidemic in Hong Kong. Journal of epidemiology and community health. 2004;58(12):988–96. Epub 2004/11/18. doi:10.1136/jech.2003.017483. PubMed PMID: 15547057; PubMed Central PMCID: PMCPMC1732647.

9. Leung GM, Lam TH, Ho LM, Ho SY, Chan BH, Wong IO, et al. The impact of community psychological responses on outbreak control for severe acute respiratory syndrome in Hong Kong. Journal of epidemiology and community health. 2003;57(11):857–63. Epub 2003/11/06. doi:10.1136/jech.57.11.857. PubMed PMID: 14600110; PubMed Central PMCID: PMCPMC1732323.

10. Huang Y, Zhao N. Generalized anxiety disorder, depressive symptoms and sleep quality during COVID-19 outbreak in China: a web-based cross-sectional survey. Psychiatry research. 2020;288:112954. Epub 2020/04/24. doi:10.1016/j.psychres.2020.112954. PubMed PMID: 32325383; PubMed Central PMCID: PMCPMC7152913.

11. Wang C, Pan R, Wan X, Tan Y, Xu L, Ho CS, et al. Immediate Psychological Responses and Associated Factors during the Initial Stage of the 2019 Coronavirus Disease (COVID-19) Epidemic among the General Population in China. International journal of environmental research and public health. 2020;17(5). Epub 2020/03/12. doi:10.3390/ijerph17051729. PubMed PMID: 32155789; PubMed Central PMCID: PMCPMC7084952.

12. Shekerdemian LS, Mahmood NR, Wolfe KK, Riggs BJ, Ross CE, McKiernan CA, et al. Characteristics and Outcomes of Children With Coronavirus Disease 2019 (COVID-19) Infection Admitted to US and Canadian Pediatric Intensive Care Units. JAMA pediatrics. 2020. Epub 2020/05/12. doi:10.1001/jamapediatrics.2020.1948. PubMed PMID: 32392288.

13. Essau CA. Frequency and patterns of mental health services utilization among adolescents with anxiety and depressive disorders. Depression and anxiety. 2005;22(3):130–7. Epub 2005/09/22. doi:10.1002/da.20115. PubMed PMID: 16175563.

14. Jongerden L, Simon E, Bodden DH, Dirksen CD, Bogels SM. Factors associated with the referral of anxious children to mental health care: the influence of family functioning, parenting, parental anxiety and child impairment. International journal of methods in psychiatric research. 2015;24(1):46–57. Epub 2014/12/17. doi:10.1002/mpr.1457. PubMed PMID: 25511424; PubMed Central PMCID: PMCPMC6878528.

15. Boman F, Stafström M, Lundin N, Moghadassi M, Törnhage CJ, Östergren PO. Comparing parent and teacher assessments of mental health in elementary school children. Scandinavian journal of public health. 2016;44(2):168–76. Epub 2015/10/30. doi:10.1177/1403494815610929. PubMed PMID: 26511589.

16. Goodman R, Ford T, Simmons H, Gatward R, Meltzer H. Using the Strengths and Difficulties Questionnaire (SDQ) to screen for child psychiatric disorders in a community sample. International review of psychiatry (Abingdon, England). 2003;15(1–2):166–72. Epub 2003/05/15. doi:10.1080/0954026021000046128. PubMed PMID: 12745328.

17. Aelterman A, Engels N, Van Petegem K, Pierre Verhaeghe J. The well-being of teachers in Flanders: the importance of a supportive school culture. Educational Studies. 2007;33(3):285–97. doi:10.1080/03055690701423085.

18. Telles S, Gupta RK, Bhardwaj AK, Singh N, Mishra P, Pal DK, et al. Increased Mental Well-Being and Reduced State Anxiety in Teachers After Participation in a Residential Yoga Program. Medical science monitor basic research. 2018;24:105–12. Epub 2018/08/01. doi:10.12659/msmbr.909200. PubMed PMID: 30061552; PubMed Central PMCID: PMCPMC6083945.

19. SurveyMonkey. San Mateo. CA.

20. Al-Rabiaah A, Temsah MH, Al-Eyadhy AA, Hasan GM, Al-Zamil F, Al-Subaie S, et al. Middle East Respiratory Syndrome-Corona Virus (MERS-CoV) associated stress among medical students at a university teaching hospital in Saudi Arabia. Journal of infection and public health. 2020. Epub 2020/02/01. doi:10.1016/j.jiph.2020.01.005. PubMed PMID: 32001194; PubMed Central PMCID: PMCPMC7102651.

21. Bults M, Beaujean DJ, Richardus JH, Voeten HA. Perceptions and behavioral responses of the general public during the 2009 influenza A (H1N1) pandemic: a systematic review. Disaster medicine and public health preparedness. 2015;9(2):207–19. Epub 2015/04/18. doi:10.1017/dmp.2014.160. PubMed PMID: 25882127.

22. DeCoster J, Gallucci M, Iselin A-MR. Best Practices for Using Median Splits, Artificial Categorization, and their Continuous Alternatives. Journal of Experimental Psychopathology. 2011;2(2):197–209. doi:10.5127/jep.008310.

23. Leung GM, Ho LM, Chan SK, Ho SY, Bacon-Shone J, Choy RY, et al. Longitudinal assessment of community psychobehavioral responses during and after the 2003 outbreak of severe acute respiratory syndrome in Hong Kong. Clinical infectious diseases : an official publication of the Infectious Diseases Society of America. 2005;40(12):1713–20. Epub 2005/05/24. doi:10.1086/429923. PubMed PMID: 15909256.

24. Spitzer RL, Kroenke K, Williams JB, Lowe B. A brief measure for assessing generalized anxiety disorder: the GAD-7. Archives of internal medicine. 2006;166(10):1092–7. Epub 2006/05/24. doi:10.1001/archinte.166.10.1092. PubMed PMID: 16717171.

25. Kroenke K, Spitzer RL, Williams JB, Monahan PO, Lowe B. Anxiety disorders in primary care: prevalence, impairment, comorbidity, and detection. Annals of internal medicine. 2007;146(5):317–25. Epub 2007/03/07. doi:10.7326/0003-4819-146-5-200703060-00004. PubMed PMID: 17339617.

26. Goodman R. The Strengths and Difficulties Questionnaire: a research note. Journal of child psychology and psychiatry, and allied disciplines. 1997;38(5):581–6. Epub 1997/07/01. doi:10.1111/j.1469-7610.1997.tb01545.x. PubMed PMID: 9255702.

27. Goodman A, Goodman R. Strengths and difficulties questionnaire as a dimensional measure of child mental health. Journal of the American Academy of Child and Adolescent Psychiatry. 2009;48(4):400–3. Epub 2009/02/27. doi:10.1097/CHI.0b013e3181985068. PubMed PMID: 19242383.

28. Silva TBF, Osório FL, Loureiro SR. SDQ: discriminative validity and diagnostic potential. 2015;6(811). doi:10.3389/fpsyg.2015.00811.

29. Goodman A. Scoring the Strengths & Difficulties Questionnaire for age 4-17. https://wwwehcapcouk/content/sites/ehcap/uploads/NewsDocuments/236/SDQEnglishUK4-17scoring-1PDF.31-5-2020.

30. Alyahri A, Goodman R. Validation of the Arabic Strengths and Difficulties Questionnaire and the Development and Well-Being Assessment. Eastern Mediterranean health journal = La revue de sante de la Mediterranee orientale = al-Majallah al-sihhiyah li-sharq al-mutawassit. 2006;12 Suppl 2:S138–46. Epub 2007/03/17. PubMed PMID: 17361685.

31. Sawaya H, Atoui M, Hamadeh A, Zeinoun P, Nahas Z. Adaptation and initial validation of the Patient Health Questionnaire - 9 (PHQ-9) and the Generalized Anxiety Disorder - 7 Questionnaire (GAD-7) in an Arabic speaking Lebanese psychiatric outpatient sample. Psychiatry research. 2016;239:245–52. Epub 2016/04/01. doi:10.1016/j.psychres.2016.03.030. PubMed PMID: 27031595.

32. SAS Institute Inc. 2011. Base SAS® 9.3 Procedures Guide. Cary NSII.

33. Lim GY, Tam WW, Lu Y, Ho CS, Zhang MW, Ho RC. Prevalence of Depression in the Community from 30 Countries between 1994 and 2014. Scientific reports. 2018;8(1):2861. Epub 2018/02/13. doi:10.1038/s41598-018-21243-x. PubMed PMID: 29434331; PubMed Central PMCID: PMCPMC5809481.

34. Löwe B, Decker O, Müller S, Brähler E, Schellberg D, Herzog W, et al. Validation and standardization of the Generalized Anxiety Disorder Screener (GAD-7) in the general population. Medical care. 2008;46(3):266–74. Epub 2008/04/05. doi:10.1097/MLR.0b013e318160d093. PubMed PMID: 18388841.

35. Borkovec TD, Ray WJ, J. S. Worry: A Cognitive Phenomenon Intimately Linked to Affective, Physiological, and Interpersonal Behavioral Processes. Cognitive Therapy and Research. 1998;22(6):561–76.

36. Paulesu E, Sambugaro E, Torti T, Danelli L, Ferri F, Scialfa G, et al. Neural correlates of worry in generalized anxiety disorder and in normal controls: a functional MRI study. Psychological medicine. 2010;40(1):117–24. Epub 2009/05/08. doi:10.1017/s0033291709005649. PubMed PMID: 19419593.

37. Sun X, So SH, Chan RCK, Chiu CD, Leung PWL. Worry and metacognitions as predictors of the development of anxiety and paranoia. Scientific reports. 2019;9(1):14723. Epub 2019/10/13. doi:10.1038/s41598-019-51280-z. PubMed PMID: 31605005; PubMed Central PMCID: PMCPMC6789003.

38. Harper CA, Satchell LP, Fido D, Latzman RD. Functional Fear Predicts Public Health Compliance in the COVID-19 Pandemic. International journal of mental health and addiction. 2020:1–14. Epub 2020/04/30. doi:10.1007/s11469-020-00281-5. PubMed PMID: 32346359; PubMed Central PMCID: PMCPMC7185265.

39. Motta Zanin G, Gentile E, Parisi A, Spasiano D. A Preliminary Evaluation of the Public Risk Perception Related to the COVID-19 Health Emergency in Italy. International journal of environmental research and public health. 2020;17(9). Epub 2020/05/01. doi:10.3390/ijerph17093024. PubMed PMID: 32349253.

40. Liao Q, Cowling BJ, Lam WW, Ng DM, Fielding R. Anxiety, worry and cognitive risk estimate in relation to protective behaviors during the 2009 influenza A/H1N1 pandemic in Hong Kong: ten cross-sectional surveys. BMC infectious diseases. 2014;14:169. Epub 2014/03/29. doi:10.1186/1471-2334-14-169. PubMed PMID: 24674239; PubMed Central PMCID: PMCPMC3986671.

41. Eisenberg SL, Eisenberg MJ. Smoking Cessation During the COVID-19 Epidemic. Nicotine & tobacco research : official journal of the Society for Research on Nicotine and Tobacco. 2020. Epub 2020/05/05. doi:10.1093/ntr/ntaa075. PubMed PMID: 32363386; PubMed Central PMCID: PMCPMC7239173.

42. Vardavas CI, Nikitara K. COVID-19 and smoking: A systematic review of the evidence. Tobacco induced diseases. 2020;18:20. Epub 2020/03/25. doi:10.18332/tid/119324. PubMed PMID: 32206052; PubMed Central PMCID: PMCPMC7083240.

43. Heerfordt C, Heerfordt IM. Has there been an increased interest in smoking cessation during the first months of the COVID-19 pandemic? A Google Trends study. Public health. 2020;183:6–7. Epub 2020/05/11. doi:10.1016/j.puhe.2020.04.012. PubMed PMID: 32388011; PubMed Central PMCID: PMCPMC7167577.

44. The COCONAL group. A future vaccination campaign against COVID-19 at risk of vaccine hesitancy and politicisation. The Lancet Infectious diseases. 2020. Epub 2020/05/24. doi:10.1016/s1473-3099(20)30426-6. PubMed PMID: 32445713; PubMed Central PMCID: PMCPMC7239623.

45. Alsuwaidi AR, Elbarazi I, Al-Hamad S, Aldhaheri R, Sheek-Hussein M, Narchi H. Vaccine hesitancy and its determinants among Arab parents: a cross-sectional survey in the United Arab Emirates. Human vaccines & immunotherapeutics. 2020:1–7. Epub 2020/05/14. doi:10.1080/21645515.2020.1753439. PubMed PMID: 32401612.

46. Harrison EA, Wu JW. Vaccine confidence in the time of COVID-19. European journal of epidemiology. 2020;35(4):325–30. Epub 2020/04/23. doi:10.1007/s10654-020-00634-3. PubMed PMID: 32318915; PubMed Central PMCID: PMCPMC7174145.

47. Al-Yateem N, Bani Issa W, Rossiter RC, Al-Shujairi A, Radwan H, Awad M, et al. Anxiety related disorders in adolescents in the United Arab Emirates: a population based cross-sectional study. BMC pediatrics. 2020;20(1):245. Epub 2020/05/27. doi:10.1186/s12887-020-02155-0. PubMed PMID: 32450837.

48. Geldsetzer P. Use of Rapid Online Surveys to Assess People’s Perceptions During Infectious Disease Outbreaks: A Cross-sectional Survey on COVID-19. Journal of medical Internet research. 2020;22(4):e18790. Epub 2020/04/03. doi:10.2196/18790. PubMed PMID: 32240094; PubMed Central PMCID: PMCPMC7124956.

